# Metagenomic sequencing identifies potential respiratory pathogens in PCR-negative subset of surveillance samples

**DOI:** 10.1101/2025.09.26.25336420

**Authors:** Anne Caroline Mascarenhas, Rose S. Kantor, James Thissen, Car Reen Kok, Monica Borucki, Christina Morales, Sharon Messenger, Crystal Jaing, Debra A. Wadford

## Abstract

Respiratory pathogens are a significant source of global morbidity, mortality, and economic burden, with the COVID-19 pandemic driving increased interest in and funding for respiratory disease surveillance. Syndromic panel multiplex nucleic acid amplification tests (NAATs) such as the BioFire Respiratory Panel (RP) are designed to identify the most common etiologic agents of respiratory illness. Untargeted metagenomic sequencing is a powerful tool for pathogen-agnostic detection, enabling the recovery of complete genomes for genomic epidemiology and variant tracking. In this study, we performed untargeted metagenomic sequencing of 305 samples previously negative by BioFire RP and SARS-CoV-2 testing and 26 samples that were previously positive by either of the diagnostic tests. A subset of 78 samples underwent probe-capture enrichment sequencing targeting human viruses. Using these methods, we identified human respiratory viruses in 16 of the 305 previously negative samples (5%). The most common viruses identified were Influenza C virus, Human Bocavirus, Rhinovirus A and C, and SARS-CoV-2. Consensus genomes were recovered for 14 viruses with >90% coverage breadth, revealing closely related Bocavirus strains from neighboring counties and distinct Rhinovirus strains across samples. We also identified 21 samples with a single predominant bacterial or fungal species in the previous negative cohort. These findings underscore the challenges of identifying causal agents from multiplex NAAT-negative cases and highlight the utility of metagenomics for expanding the scope of pathogen surveillance.

## 1. Introduction

Respiratory pathogens cause significant morbidity, mortality, and economic burden with an estimated 12.8 billion new upper respiratory infections globally in the year 2021 ^1^. The COVID-19 pandemic led to increased interest in and funding for the surveillance of respiratory diseases. This surveillance has primarily been conducted via qPCR and PCR-based panels for clinical specimens, with a focus on SARS-CoV-2, influenza A virus, and respiratory syncytial virus (RSV) ^2-4^. Wastewater-based surveillance has also provided a partial view of circulating pathogens, although some respiratory pathogens may not be shed into wastewater at detectable concentrations ^5-7^.^3^

While clinical surveillance increased during and after the pandemic, the range of targets has generally been limited to commonly occurring pathogens whose genomes are conserved enough to be detectable by established PCR methods. Hence, rarer or more divergent pathogens may have been missed. Separately, use of pathogen-agnostic untargeted metagenomic and metatranscriptomic sequencing has increased for characterization of respiratory infections in clinical samples ^8-10^. Recently, several groups have developed validated clinical metagenomic workflows for respiratory viruses ^11,12^. Additionally, commercial probe-capture sequencing panels for broad-range viral enrichment, including respiratory viruses, have recently been shown to increase the sensitivity of sequencing ^13,14^. Clinical metagenomic sequencing may even produce complete pathogen genomes, which, as demonstrated for SARS-CoV-2, can be useful for variant tracking, for genomic epidemiology and for predicting vaccine and treatment efficacy ^15^. Yet despite its potential utility, metagenomics has not been widely applied in surveillance beyond SARS-CoV-2 due to its relatively high cost.

During the COVID-19 pandemic, the California SARS-CoV-2 and Respiratory Virus Surveillance System (CalSRVSS) was established by the California Department of Public Health (CDPH) to provide outpatient testing of symptomatic individuals in 10 counties across California^16^. From 2020-2023, this program collected 15,325 samples from symptomatic individuals, which were tested for SARS-CoV-2 and 22 additional pathogens included on the BioFire Respiratory Pathogens panel. Of these, 55% were negative for all 22 BioFire pathogen analytes (unpublished data). These BioFire- and SARS-CoV-2 negative samples provided a unique opportunity to examine whether less commonly recognized pathogens may have been associated with the respiratory symptoms observed from these cases. We randomly selected 305 previously negative samples and 26 previously positive samples from the CalSRVSS sample set for untargeted sequencing and analyzed a subset of 78 samples using semi-targeted metagenomics. Here, we report on the frequency of potential pathogens detected.

## 2. Methods

### Sample collection

De-identified residual nasopharyngeal and other swab eluate samples (n=305, identifiers “N#”) from adult and pediatric patients with upper respiratory tract infection symptoms were obtained from the California Department of Public Health (CDPH) as part of the California SARS-CoV-2 and Respiratory Virus Surveillance System (CalSRVSS) initiative (**Table S1**)^16^. These samples were collected during outpatient visits between July 2020 to January 2022 in 10 counties in California and were negative for SARS-CoV-2 and for 18 viruses and 4 bacteria included in the BioFire Respiratory Panel 2.0 or 2.1 (https://www.biofiredx.com), which covers pathogens commonly associated with upper respiratory tract infections^17^. A set of known positive swabs including SARS-CoV-2 positive samples from individuals previously infected with SARS-CoV-2 (“reinfections”, n=7) and BioFire positive samples (n=19) were included for comparison (identifiers “P#”; **Table S1**). Samples were received in PrimeStore Molecular Transport Medium (ThermoFisher Scientific, Waltham, MA). The sample IDs do not reflect patient identifiers and were solely used for sample tracking purposes to protect the patient ID.

The CalSRVSS project was reviewed by the State of California Health and Human Services Agency Committee for the Protection of Human Subjects and determined to be considered Not Research or Exempt (Project # 2021-168).

### Total nucleic acid extraction

Total nucleic acid (DNA and RNA) was extracted from 500 μL of each eluate sample as follows. First, cells were lysed in 100μL ATL Buffer (Qiagen, Redwood City, CA) with bead-beating for 40s at 6 m/s with FastPrep-24 5G equipment using lysing matrix F tubes (MP Biomedicals, Burlingame, CA). Total nucleic acid was extracted from 200 μL of the resulting lysate using the IndiSpin Pathogen Kit (Indical Bioscience, Leipzig, Germany) following the manufacturer’s protocol with final elution in 55μL AVE Buffer. DNA and RNA were quantified separately using the Qubit High Sensitivity dsDNA and RNA kits, respectively, on a Qubit 3.0 fluorometer (Invitrogen, Carlsbad, CA) using the recommended instructions.

### cDNA synthesis and whole genome amplification

The first strand cDNA synthesis reaction contained 11μL of sample, 1μL of random hexamer primers (50mM; Integrated DNA Technologies, Coralville, IA) and 1μL of dNTP mix (10mM; Thermo Fisher Scientific, Waltham, MA) and was incubated at 65°C for 5 min, and 5 min on ice-water slurry. Synthesis was completed by addition of the remaining reaction components (4 μL of 5x First Strand Synthesis Buffer, 1 μL of 0.1 M DTT, 1 μL of RNAseOUT, 1 μL of SuperScript IV reverse transcriptase and 1 μL of DEPC water (Thermo Fisher Scientific, Waltham, MA) to the cooled samples followed by incubation in a thermocycler (protocol: 23°C for 10 min, 55°C for 60 min, and 80°C for 10 min). A total of 10μL from each DNA/cDNA sample was amplified using the Whole Transcriptome Amplification kit (Qiagen, Redwood City, CA) following the manufacturer’s guidelines. Amplified samples were purified with AMPure XP magnetic beads (Beckman Coulter, Brea, CA) using a slightly modified version of the recommended protocol: 1.8x AMPure XP magnetic beads, 3x Ethanol 70% washes, and elution in 60 μL of Ultra-Pure Water (UPW). Purified samples were used in downstream library preparation protocols for sequencing.

### Untargeted metagenomic sequencing

The untargeted metagenomics DNA libraries were prepared from 200-300 ng of amplified nucleic acid with the Illumina DNA Prep kit (Illumina, San Diego, CA) following the standard manufacturer’s protocol but using the Illumina UD 10-bases Nextera Indexes from IDT with 5 PCR cycles and 30μL of RSB for final library elution. Final libraries were quantified, and quality was assessed with the High Sensitivity D5000 assay on a TapeStation 4200 instrument (Agilent Technologies, Santa Clara, CA). Libraries were pooled in equimolar quantities for multiplexing and sequenced in 300 cycles, with 2x151bp inserts and 10bp indexes in paired-end mode on an Illumina NextSeq2000 using its on-board denaturing function.

### Twist CVRP library preparation and sequencing

Samples that were selected for enrichment with the Twist comprehensive viral research panel (Twist Bioscience, South San Francisco, CA) were prepared as follows. First, DNA libraries from the whole genome amplified samples were prepared with the Twist Total Nucleic Acids Library Preparation EF Kit 2.0 for Viral Pathogen Detection and Characterization starting from Step 3 “Perform DNA fragmentation, end repair and dA-tailing” of the protocol. The whole-genome amplified samples (1-2 ng of in 25μL of UPW) were fragmented, repaired and dA-tailed under the following thermal cycler conditions: 1) hold at 4°C, 2) incubation at 37°C for 20 min to generate 180-220bp inserts, 3) 65°C for 30 min and 4) hold at 4°C. Then, the Twist universal adapters were ligated to the dA-tailed DNA fragments, and libraries were purified with magnetic beads, as per the instructions. The adapter-ligated libraries were amplified with the Twist UDI Primers (10bp in length) using 10 cycles for the Polymerase Reaction (PCR), purified using the manufacturer’s instructions but eluted in 25μL of UPW. The resulting libraries were quantified with the High Sensitivity dsDNA Kit on a Qubit 2.0 fluorometer.

Libraries were enriched with the Twist Comprehensive Viral Research Panel (CVRP) (36.8 Mb) following the Twist Target Enrichment Standard Hybridization v2 Protocol. Briefly, libraries were pooled in equal quantities (187.5ng each) in groups of 8 samples for a multiplexed 16h hybridization with the probes in the enrichment panel. The hybridized targets were captured with streptavidin beads, amplified using a panel size of 10-50Mb using 8 PCR cycles for multiplexed samples, followed by a purification step with DNA Purification Beads and eluted in 35μL of UPW. The viral enriched libraries were quantified with a High Sensitivity dsDNA Kit on a Qubit 2.0, and quality was measured using a High Sensitivity D5000 assay on a TapeStation 4200 instrument. The Twist library pools were multiplexed in equal quantities and sequenced with 300x cycles with 2x 151bp inserts and 10bp indexes in paired-end mode on an Illumina NextSeq2000 using its on-board denaturing function.

### Dataset pre-processing

Illumina paired-end metagenomic sequencing reads were demultiplexed and converted from bcl to fastq file format with bcl2fastq v2.20.0.422 (Illumina, San Diego, CA) using default parameters. Reads were filtered and trimmed with Fastp v0.23.4 ^18,19^ using the command line switches ‘-y -g --poly_g_min_len=5 -x -- poly_x_min_len=5’. Human genomic content was removed from the metagenomic datasets by mapping the sequencing reads to the reference human genome (GRCh38.p14; RefSeq Assembly: GCF_000001405.40) using minimap2 v2.26-r1175 ^20^ configured for short-read alignment with ‘-x sr’. The human-depleted sam files were converted to bam with Samtools v1.13 ^21,22^ using the view algorithm and the command line options ‘-bSh -f 4’. Decontaminated bam files were sorted and converted to fastq with samtools.

All of the paired-end sequencing reads for untargeted and Twist-enriched metagenomic datasets were de-identified as recommended by the National Center of Biotechnology Information (NCBI) prior to deposition in the Sequence Read Archive (SRA) ^23^. Briefly, Illumina paired-end reads were interleaved with SeqFu v1.22.3 ^24^ using the built-in interleave function and careful mode enabled (-c). Interleaved datasets were de-identified using the NCBI’s Human Read Removal Tool (HRRT) v2.2.1 tool (SRA Human Scrubber; https://github.com/ncbi/sra-human-scrubber)^25^ with the ‘-x’ command line switch. The ‘scrubbed’ reads were de-interleaved with SeqFu using its deinterleave function and careful mode enabled (-c).

### Taxonomic classification and analysis

The filtered metagenomic reads were taxonomically classified using a Lower Common Ancestor (LCA) approach by Centrifuge v1.0.4 ^26^ through read alignment against the NCBI nucleotide (NT) database ^27^ curated in January 2023 ^28^. Centrifuge was configured with a minimum hit length of 15 and best-hit reporting using ‘--min-hitlen 15’ and ‘-k 1’. Results were refined with Recentrifuge v1.12.0^29^ using a minimum hit length of 40. Recentrifuge read count data was imported into R+ v4.2.0 and Python v3.13.0 (pandas v2.2.3) for downstream microbiome (bacterial, fungal, and viral) analyses. Within R, taxonomic compositional analysis was carried out using the phyloseq package v1.48.0 ^30^. Species-level hits were cross-referenced with a curated list of human infectious pathogens compiled from the Swiss Institute of Bioinformatics ViralZone database (De Castro et al., 2024), the NCBI Pathogen Detection database (Information, 2016) and two previously published compilations of bacterial (Bartlett et al., 2022) and fungal (Bartoszewicz et al., 2022) pathogens. Excluded from consideration were non-respiratory viruses including polyomaviruses, papillomaviruses, herpesviruses, erythroviruses, Molluscum contagiosum virus, GB virus C, rotaviruses, Torque teno virus, and picobirnaviruses (**Supplementary Figure S2**). We also removed pervasive low read-count hits to human immunodeficiency virus (HIV) caused by human-contaminated viral genomes in the database. Viral detection in previously negative and positive samples used a threshold of ≥10 reads per virus based on Recentrifuge. Two no-template control libraries were also sequenced and found to contain no reads hitting to any human pathogenic virus. Heatmaps were generated showing species-level read counts (with the exception of SARS-CoV-2, which was “no rank” within the taxonomy database used), using plotnine v0.13.6.

### Viral genome analysis

For samples found by Recentrifuge to contain ≥ 10 reads hitting to Bocavirus, Rhinovirus A, Rhinovirus C, Influenza C virus, or SARS-CoV-2, additional analysis was conducted. First, all complete genomes classified as Bocaparvovirus (taxID 1507401), Rhinovirus A (taxID 147711), and Rhinovirus C (taxID 463676) were downloaded from Genbank using the NCBI Virus portal (https://www.ncbi.nlm.nih.gov/labs/virus/vssi/#/; accessed April 7, 2025). For each of these groups, Genbank genomes were clustered at 95% identity and 75% coverage using mmseqs2 ^31^ (version 18d8ddc2f468b8a00cec9addb3d2328eb790ac9e). Reads from each sample containing each virus were mapped to cluster representatives, using minimap2 (v2.28-r1209) ^20^ without secondary hits. Mapping results were filtered to remove supplementary hits (samtools view -F 0x800), sorted, indexed, and used as input for coverage calculations by samtools (v1.21). The genome with highest coverage breadth for each virus in each sample was used for a final mapping step, which provided coverage information for untargeted and targeted sequencing (**Table 1**). Masked consensus genomes were extracted using samtools (v1.21) with default settings (requiring a frequency of 0.75 to make a base call) from the final mapping results using targeted sequencing data. Best BLAST hits were identified using BLASTn against the NCBI core-nt database (accessed April 14, 2025). For influenza C virus and SARS-CoV-2, the RefSeq entries (strains C/Ann Arbor/1/50 and Wuhan Hu-1, respectively) were used as the reference genomes in a single mapping step followed by coverage calculations and extraction of the consensus. Mapping achieved near 100% coverage breadth for most samples, suggesting these reference genomes were acceptable starting points. For SARS-CoV-2, variant lineages were identified from consensus sequences using the Nextclade web interface (https://clades.nextstrain.org/ accessed April 28, 2025).

**Table 1.**
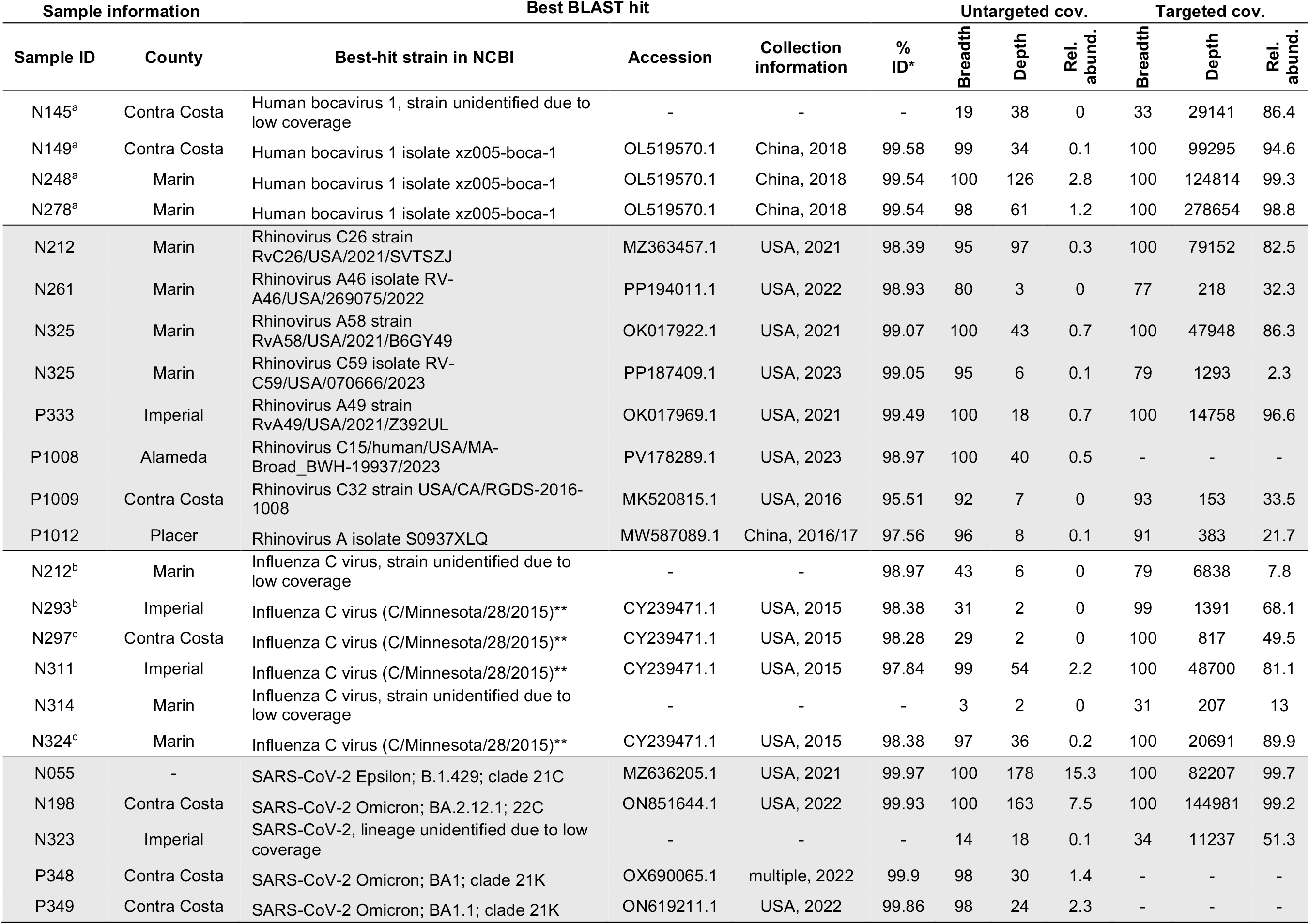
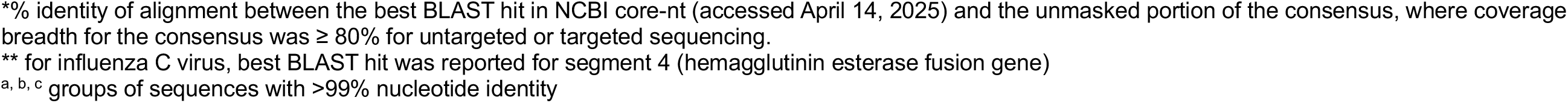
Identification, coverage, and relative abundance of viral consensus genomes recovered.

| Sample information |  | Best BLAST hit |  |  |  | Untargeted cov. |  |  | Targeted cov. |  |  |
| --- | --- | --- | --- | --- | --- | --- | --- | --- | --- | --- | --- |
| Sample ID | County | Best-hit strain in NCBI | Accession | Collection information | % ID* | Breadth | Depth | Rel. abund. | Breadth | Depth | Rel. abund. |
| N145<sup>a</sup> | Contra Costa | Human bocavirus 1, strain unidentified due to low coverage | - | - | - | 19 | 38 | 0 | 33 | 29141 | 86.4 |
| N149<sup>a</sup> | Contra Costa | Human bocavirus 1 isolate xz005-boca-1 | OL519570.1 | China, 2018 | 99.58 | 99 | 34 | 0.1 | 100 | 99295 | 94.6 |
| N248<sup>a</sup> | Marin | Human bocavirus 1 isolate xz005-boca-1 | OL519570.1 | China, 2018 | 99.54 | 100 | 126 | 2.8 | 100 | 124814 | 99.3 |
| N278<sup>a</sup> | Marin | Human bocavirus 1 isolate xz005-boca-1 | OL519570.1 | China, 2018 | 99.54 | 98 | 61 | 1.2 | 100 | 278654 | 98.8 |
| N212 | Marin | Rhinovirus C26 strain RvC26/USA/2021/SVTSZJ | MZ363457.1 | USA, 2021 | 98.39 | 95 | 97 | 0.3 | 100 | 79152 | 82.5 |
| N261 | Marin | Rhinovirus A46 isolate RV-A46/USA/269075/2022 | PP194011.1 | USA, 2022 | 98.93 | 80 | 3 | 0 | 77 | 218 | 32.3 |
| N325 | Marin | Rhinovirus A58 strain RvA58/USA/2021/B6GY49 | OK017922.1 | USA, 2021 | 99.07 | 100 | 43 | 0.7 | 100 | 47948 | 86.3 |
| N325 | Marin | Rhinovirus C59 isolate RV-C59/USA/070666/2023 | PP187409.1 | USA, 2023 | 99.05 | 95 | 6 | 0.1 | 79 | 1293 | 2.3 |
| P333 | Imperial | Rhinovirus A49 strain RvA49/USA/2021/Z392UL | OK017969.1 | USA, 2021 | 99.49 | 100 | 18 | 0.7 | 100 | 14758 | 96.6 |
| P1008 | Alameda | Rhinovirus C15/human/USA/MA-Broad_BWH-19937/2023 | PV178289.1 | USA, 2023 | 98.97 | 100 | 40 | 0.5 | - | - | - |
| P1009 | Contra Costa | Rhinovirus C32 strain USA/CA/RGDS-2016-1008 | MK520815.1 | USA, 2016 | 95.51 | 92 | 7 | 0 | 93 | 153 | 33.5 |
| P1012 | Placer | Rhinovirus A isolate S0937XLQ | MW587089.1 | China, 2016/17 | 97.56 | 96 | 8 | 0.1 | 91 | 383 | 21.7 |
| N212<sup>b</sup> | Marin | Influenza C virus, strain unidentified due to low coverage | - | - | 98.97 | 43 | 6 | 0 | 79 | 6838 | 7.8 |
| N293<sup>b</sup> | Imperial | Influenza C virus (C/Minnesota/28/2015)** | CY239471.1 | USA, 2015 | 98.38 | 31 | 2 | 0 | 99 | 1391 | 68.1 |
| N297<sup>c</sup> | Contra Costa | Influenza C virus (C/Minnesota/28/2015)** | CY239471.1 | USA, 2015 | 98.28 | 29 | 2 | 0 | 100 | 817 | 49.5 |
| N311 | Imperial | Influenza C virus (C/Minnesota/28/2015)** | CY239471.1 | USA, 2015 | 97.84 | 99 | 54 | 2.2 | 100 | 48700 | 81.1 |
| N314 | Marin | Influenza C virus, strain unidentified due to low coverage | - | - | - | 3 | 2 | 0 | 31 | 207 | 13 |
| N324<sup>c</sup> | Marin | Influenza C virus (C/Minnesota/28/2015)** | CY239471.1 | USA, 2015 | 98.38 | 97 | 36 | 0.2 | 100 | 20691 | 89.9 |
| N055 | - | SARS-CoV-2 Epsilon; B.1.429; clade 21C | MZ636205.1 | USA, 2021 | 99.97 | 100 | 178 | 15.3 | 100 | 82207 | 99.7 |
| N198 | Contra Costa | SARS-CoV-2 Omicron; BA.2.12.1; 22C | ON851644.1 | USA, 2022 | 99.93 | 100 | 163 | 7.5 | 100 | 144981 | 99.2 |
| N323 | Imperial | SARS-CoV-2, lineage unidentified due to low coverage | - | - | - | 14 | 18 | 0.1 | 34 | 11237 | 51.3 |
| P348 | Contra Costa | SARS-CoV-2 Omicron; BA1; clade 21K | OX690065.1 | multiple, 2022 | 99.9 | 98 | 30 | 1.4 | - | - | - |
| P349 | Contra Costa | SARS-CoV-2 Omicron; BA.1.1; clade 21K | ON619211.1 | USA, 2022 | 99.86 | 98 | 24 | 2.3 | - | - | - |

\*% identity of alignment between the best BLAST hit in NCBI core-nt (accessed April 14, 2025) and the unmasked portion of the consensus, where coverage breadth for the consensus was $\geq 80\%$ for untargeted or targeted sequencing. \*\* for influenza C virus, best BLAST hit was reported for segment 4 (hemagglutinin esterase fusion gene) a, b, c groups of sequences with $>99\%$ nucleotide identity

## 3. Results

We randomly selected 305 BioFire-negative samples for sequencing to detect less common respiratory pathogens not included as part of the BioFire Respiratory Pathogens panel. Samples were collected from individuals with respiratory symptoms as part of CalSRVSS across five counties in California between July 2020 and December 2022 ^16^. In parallel, a set of known viral positive samples and SARS-CoV-2 reinfection samples was selected based on positive results for one or more respiratory pathogens (“previous positives”, n=26). Untargeted Illumina sequencing produced an average of 10.8 million reads per sample, and probe-capture enrichment (Twist CVRP) on a subset of 78 samples (56 previous negative, 22 previous positives) produced an average of 4.9 million reads per sample (**Supplementary Table S1**). In nearly all samples, a majority of untargeted sequencing reads were derived from the human host, with a few exceptions in which bacteria dominated (**Supplementary Figure S1**). After host read removal, untargeted sequence data were dominated by unclassified sequences and bacterial sequences. Meanwhile, targeted sequencing via probe-capture enrichment using the Twist CVRP yielded a relatively high fraction of viral reads for a subset of previous negative and most previous positive samples **(Figure 1A**).

**Figure 1.**
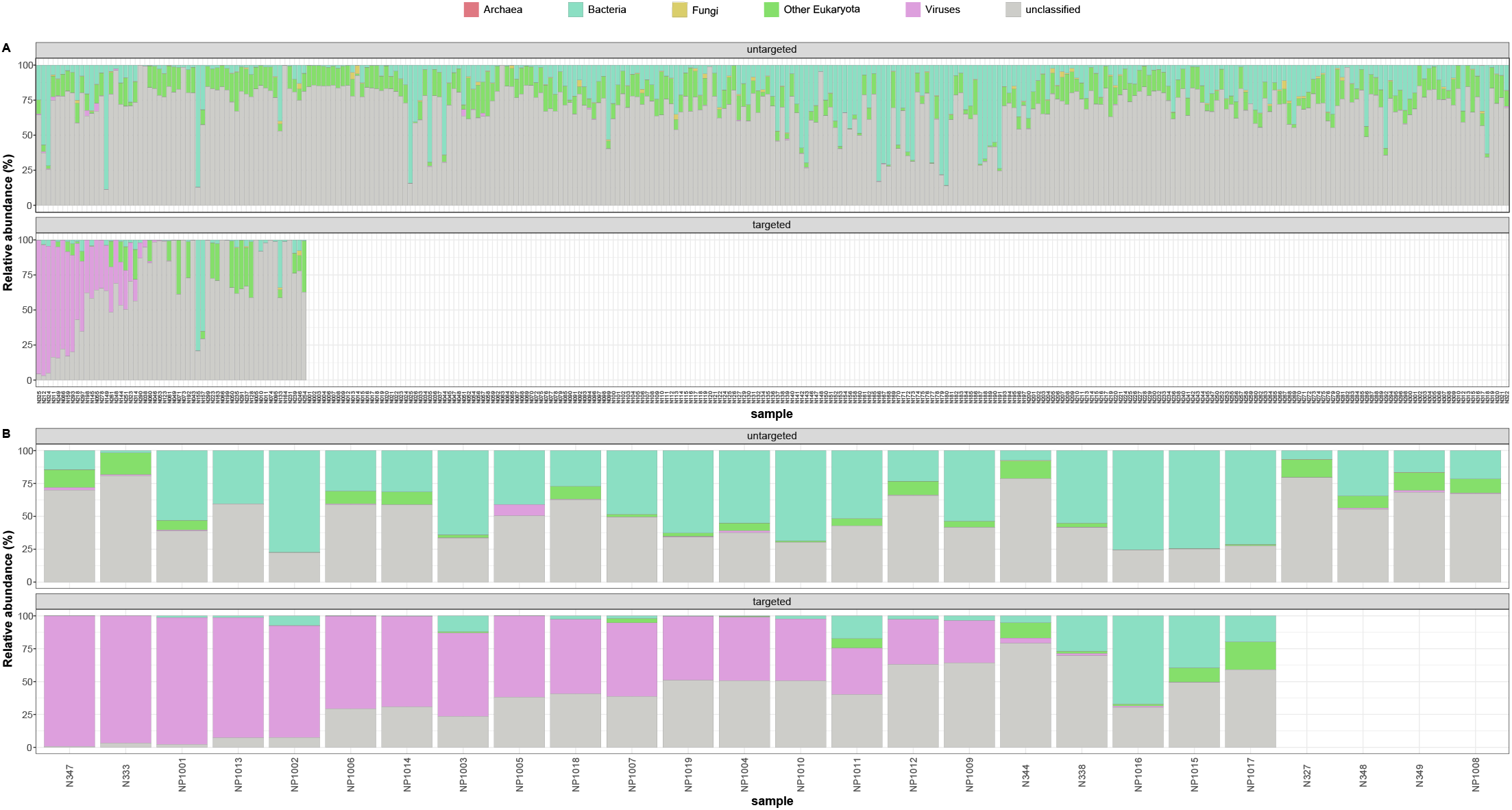
Relative abundance of viral, archaeal, bacterial, fungal, non-human and non-fungal eukaryotic reads across the data cohort. Previous (**A**) positive and (**B**) negative samples were sequenced with untargeted metagenomics (top panel), while a subset also underwent viral enrichment (bottom panel). Data are presented after human host removal (see **Supplementary Figure S1** for inclusion of host data) and taxonomy classification with Centrifuge and Recentrifuge.

### 3.1 Viral pathogen detection

We first employed Centrifuge and Recentrifuge as a highly sensitive method for detecting viral reads, cross-referencing hits against a list of human pathogenic viruses. In previous positive samples, expected viruses were largely confirmed by both untargeted and probe-capture sequencing, with some exceptions (**Figure 2A**). In particular, of 7 SARS-CoV-2 previous positive samples, just two were successfully sequenced with untargeted sequencing. The remaining 5 samples underwent targeted sequencing to look for additional infections. One of these had a known co-infection with human coronavirus 299E, which was confirmed by sequencing while SARS-CoV-2 was not detected. Rhinovirus was detected in one other SARS-CoV-2 non-detect sample with probe-capture enrichment. Expected viruses were not detected in three other previous positives even with targeted sequencing **(Figure 2A)**.

**Figure 2.**
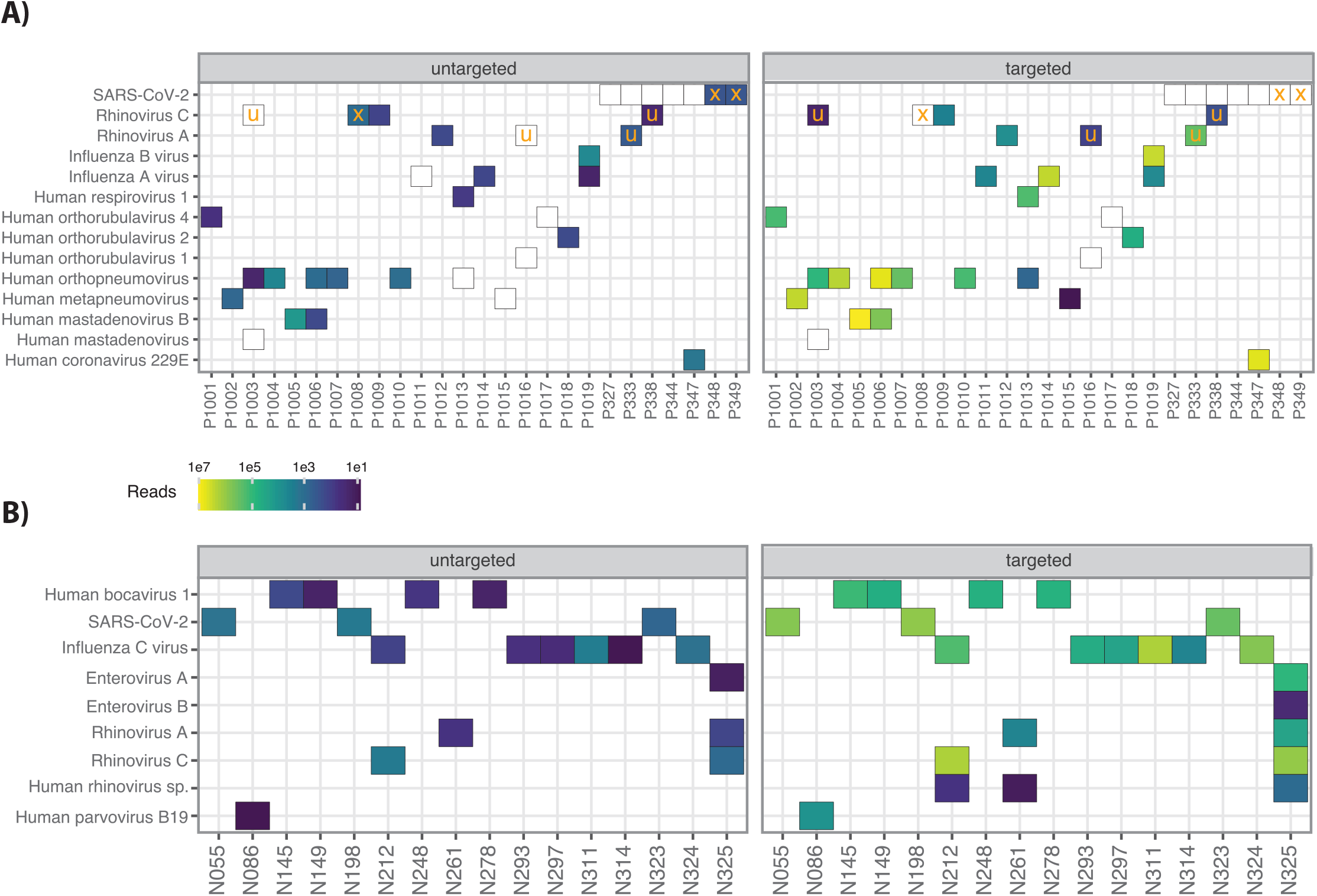
Human respiratory virus detection. Detection in (A) previous positive samples and (B) previous negative samples shows substantial improvement with probe-capture enrichment (right). Fill color indicates read counts based on Recentrifuge, shown where ≥ 10 reads per virus, per sample; note log10 scale. Samples that did not undergo probe-capture enrichment are indicated with “x”, while unexpected viral detections are indicated with “u”. White tiles for previous positive samples indicate viruses expected based on previous results that were not detected by sequencing. Note that some samples contained multiple viruses.

Human respiratory viruses were detected in16 of 305 (5%) of previous negative samples via untargeted and targeted sequencing. Probe-capture enrichment improved viral sequence recovery relative to untargeted sequencing (**Figure 2B**). The most commonly detected viruses in previous negative samples were Influenza C virus (6 samples), Human Bocavirus (4 samples), Rhinovirus A and C (3 samples), and SARS-CoV-2 (3 samples). One additional sample contained Human parvovirus B19 (**Figure 2B**).

To assess genomic novelty and potential strain-level relationships between the most common viruses in previous negative samples, we mapped untargeted and targeted sequencing reads to corresponding reference sequences (see Methods). Consensus genomes were extracted for 14 viruses with > 90% coverage breadth. We recovered 3 complete genomes and one partial genome for Human Bocavirus 1 from samples collected from the nearby counties of Contra Costa and Marin between 2021-2022 (**Table 1**). All genomes were closely related, differing from one another at a total of 25 nucleotide positions (99.6% identity). Sequences were also closely related to the nearest GenBank reference (99.5% identity).

We detected Rhinovirus A (RhV-A) from two previous negative samples (N261 and N325; **Figure 2B**), one known positive sample (P1012), and unexpectedly from two samples previously positive for other viruses (P1016 and P333; **Figure 2A**). Rhinovirus C (RhV-C) was identified from two previous negative samples (N212 and N325; **Figure 2B**), two previous positive samples (P1008 and P1009), and one SARS-CoV-2 positive sample using Centrifuge. From these samples, we recovered 7 RhV genomes with > 90% coverage breadth and one partial RhV genome. Notably, each sample contained distinct RhV strain(s), and the percent identity to the nearest reference genome ranged from 95.5% to 99.5%. One sample (N325) contained a co-infection of Rhinovirus A58 and C59, of which both sequences were recovered (**Figure 2B, Table 1**). This sample also contained Enterovirus A, according to Centrifuge-based analysis, although this was not analyzed further as no other samples contained Enterovirus A. We also detected a co-infection in a previous negative sample (N212), which had both RhV-C and Influenza C virus (**Figure 2B**).

From the six samples in which Influenza C virus was identified, we recovered 4 near-complete (>99% coverage breadth) and 2 partial viral genomes. BLAST results against NCBI core-nt for the consensus sequences of the hemagglutinin esterase fusion gene (HEF), suggested that the four near-complete genomes are likely most closely related to Influenza C virus strain C/Minnesota/28/2015. To assess the relationship between Influenza C virus genomes, we aligned the consensus sequences of the hemagglutinin esterase fusion (HEF) and M segments. This alignment showed two sequence groups, each with >99% within-group identity (**Table 1**). All samples positive for Influenza C were collected between February and July 2022 and represented 3 different counties. However, the near-identical sequences in each group were not from the same county (**Table 1**).

The presence of SARS-CoV-2 in three previous negative samples was unexpected. We recovered complete genomes from two of these samples and were able to classify these as belonging to the Epsilon (sample N055) and Omicron BA.2 (sample N198) lineages, respectively (**Table 1)**. We searched the two complete and one partial SARS-CoV-2 genomes for the primer and probe sequences of the CDC RT-PCR diagnostic assay for SARS-CoV-2 ^32^. For one genome (N055), primer and probe sequences for targets N1, N2, and N3 were perfect matches, and for the other two genomes (samples N198 and N323), the primer and probe sequences for N3 were exact matches while the N1 target had a single mismatch near the 5’ end of the probe. Genomes were also recovered from two previous positive samples, allowing classification of these as Omicron BA.1 and BA1.1 (**Table 1)**.

Use of probe-capture enrichment improved genome coverage by as much as 71%, producing the biggest gains for influenza C virus (**Table 1**). For three Rhinovirus genomes, coverage breadth decreased after probe-capture enrichment although the fraction of total reads mapped to this genome (relative abundance) increased.

### 3.2 Bacterial and fungal pathogens

Following viral assessment, we screened the 305 previous negative samples to identify those in which a single bacterial or fungal species was detected at ≥50% relative abundance after host-read removal (**Figure 3**). Twenty samples had high prevalence of a single bacterial species, such as *Dolosigranulum pigrum, Moraxella* spp., *Staphylococcus epidermidis* and *Acinetobacter junii*. Surprisingly, 10 samples with high bacterial content were dominated by *Pseudomonas azotoformans*. Additionally, four bacteria-dominated samples were also positive for viral human pathogens, suggesting possible viral and bacterial co-infections. Lastly, a single sample (N014) was found to be dominated by fungi, with a fungal relative abundance of 72%, most of which were classified as *Penicillium* and *Aspergillus* spp. (**Figure 3**).

**Figure 3.**
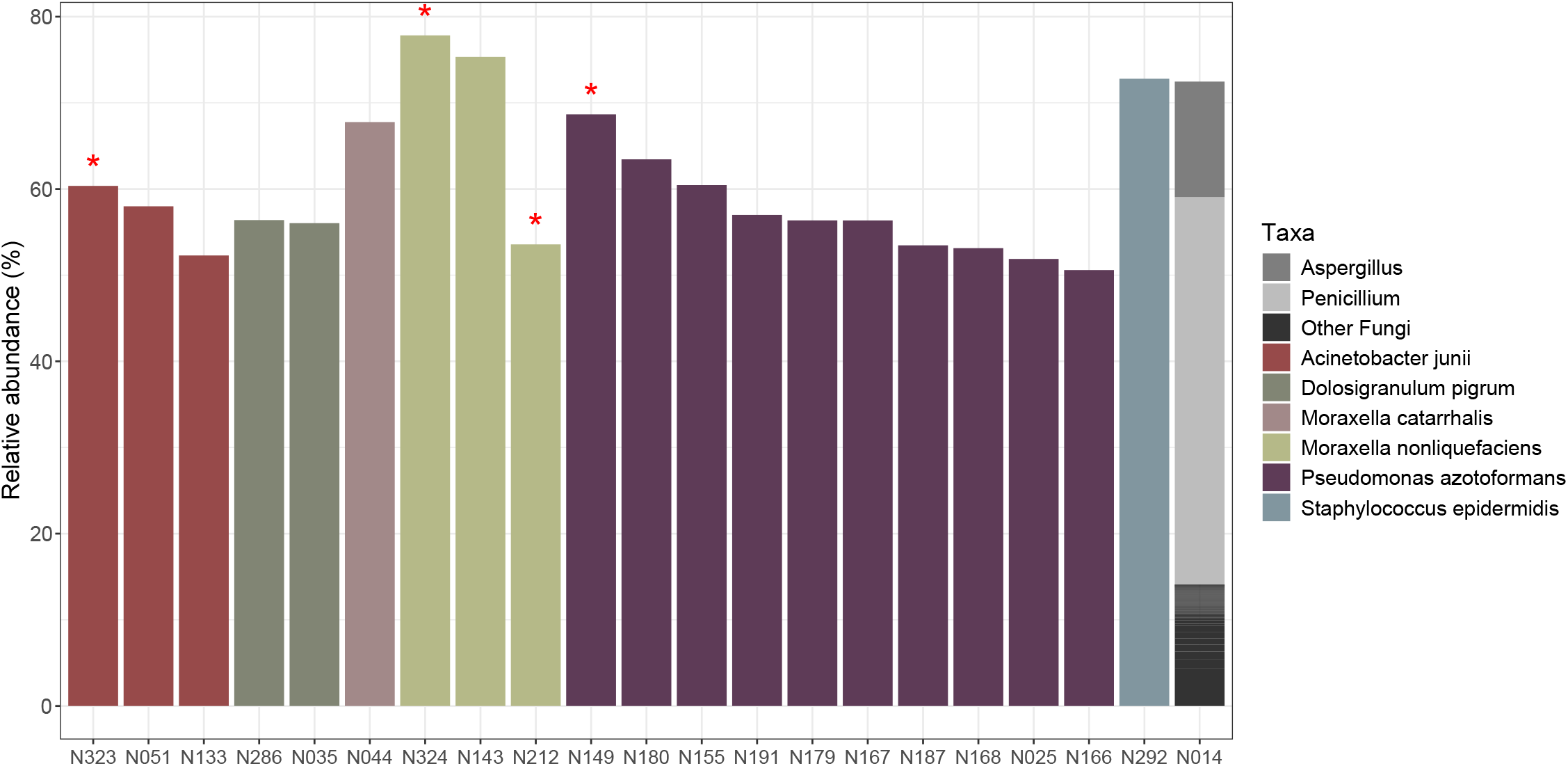
Previous negative samples dominated by a single bacterial or fungal species. Only samples with bacterial or fungal relative abundance ≥ 50% considered dominated by these signals. Red asterisks indicate samples in which respiratory viruses were also detected.

## 4. Discussion

### 4.1 Pathogen detection and limitations

Our metagenomic analysis identified Influenza C virus and Human Bocavirus 1 from previous negative samples. Recent epidemiological studies have shown that these viruses are relatively common, especially in children ^33^, yet are not included in routine test respiratory panels. Our results suggest that both Influenza C virus and Human Bocavirus 1 circulated broadly and locally within California, based on closely related genomes of each virus and counties where the respective samples were collected. The Human Bocavirus 1 detections should be interpreted with caution as this virus is often shed well after acute infection ^34^ and has been detected in healthy controls ^35^, and as a common coinfection. Notably, we did not identify any other viruses in these samples, and recent work has shown that Bocavirus may cause respiratory symptoms as a single infection.

In addition to identifying less common viral pathogens by metagenomics, we also detected bacterial and fungal colonizers. Twenty samples were dominated by a single bacterial species, which included both opportunistic pathogens and environmental microorganisms **(Figure 3**). Specifically, four samples contained high relative abundances of *Moraxella catarrhalis* or *Moraxella nonliquefaciens* ^36,37^. These are opportunistic pathogens but are also common members of the respiratory tract microbiome, making it difficult to interpret the significance of this finding without additional data on absolute abundance and clinical symptoms. Species such as *Acinetobacter junii* and *Pseudomonas azotoformans* were also predominant in some samples. As these species are not commonly linked to human respiratory infections or to the healthy respiratory microbiome, we speculate that their high abundance in some samples may reflect transient colonization. Likewise, the high relative abundance of fungi in one sample was surprising, but without additional data it remains unclear whether this represented an infection. Lastly, we observed instances of potential viral and bacterial co-infection. Previous studies have described the association of secondary bacterial infections during respiratory viral illness with increased disease severity and mortality ^38,39^. This is potentially important, as microbial community composition is thought to play a role in disease progression and outcome of respiratory viral infections ^40^.

Overall, by metagenomic sequencing of 305 previous negative samples, we were able to identify 16 (5%) viral and 35 (12%) bacterial/fungal pathogens. While it is unclear whether these detections were from outright infections or part of the normal microbiome, for bacterial detections in particular, the overwhelming majority (254/305 or 83%) of the previous negative samples from individuals who were symptomatic for respiratory illness yielded no detections of known pathogens. Possible limitations for detection in this study include: 1) non-infectious etiology of respiratory symptoms (e.g., asthma or allergy), 2) poor/low quality of sample/nucleic acid due to multiple freeze-thaw events, 3) low concentration of pathogen nucleic acid, 4) improper timing of sample collection, 5) improper sample collection technique, and/or 6) high concentration of host nucleic acid material that may have interfered with pathogen sequencing reactions.

### 4.2 Methods evaluation

We observed several discrepancies between sequencing results and the prior surveillance test results from BioFire and SARS-CoV-2 diagnostic assays. Of 31 expected viruses from previous positive samples, 8 were not detected by either untargeted or targeted sequencing. In these instances, the sensitivity of sequencing may have been limited due to long sample storage times (up to 43 months), freeze-thaw cycles between previous testing and sequencing, or low initial concentration of viral nucleic acids in the samples. It is also possible that the previous results were false-positive. Additionally, we identified unexpected positive samples containing SARS-CoV-2. Our sequence-based analysis indicated that these viral sequences could have been detected by the CDC RT-qPCR assay, and we are uncertain as to why SARS-CoV-2 was not detected by PCR. Separately, from previously reported negative specimens, we unexpectedly detected RhV genomes (RhV-A and RhV-C) and SARS-CoV-2 genomes via sequencing (**Figure 2**). The RhVs we detected from previous negative samples were likely too divergent to be captured by the BioFire panel. In particular, one sample contained a sequence closely related to the novel Rhinovirus C59 genotype recently reported from western Washington state, USA (**Table 1**) (30).

As expected, probe-capture enrichment generally increased the coverage breadth and depth of target genomes (**Table 1**). However, enrichment did not always recover complete genomes even when relative abundance of target reads was >50% (e.g. samples N145 and N323), suggesting selective degradation of the target genome in the sample or incomplete capture by probes due to sequence divergence. Notably, coverage breadth for RhV genomes was lower in targeted than untargeted sequencing, suggesting the probe panel, like the BioFire panel, had limited representation of this diverse viral group.

## 5. Conclusion

Our results support that for public health surveillance, metagenomic sequencing can provide a complementary approach to common clinical diagnostic tools. Knowledge about the prevalence of rarer and more divergent viruses in this dataset and from future sequencing-based surveillance efforts could aid clinical and public health practitioners in expanding test panels for routine diagnostics, surveillance, and outbreak investigations. Untargeted metagenomic sequencing can provide a truly pathogen agnostic method of surveillance, as compared to targeted NAAT based assays. However, more work is needed to determine the appropriate depth of sequencing necessary to identify the rarer and more divergent viruses. Ultra deep sequencing may be cost-prohibitive for analysis of large quantities of samples. The use of a broad probe-capture panel prior to metagenomic sequencing represents an intermediate and potentially more cost-effective option to recover near-complete viral genomes for downstream analysis, as we have demonstrated in this study. In summary, we found that a multi-pronged approach combining NAAT-based test panels with unbiased and targeted sequencing was successful to identify both common and uncommon respiratory pathogens. Pathogen-agnostic sequencing is an active research area that has made significant progress over the past few years. This capability will advance pathogen surveillance and diagnostics, shorten time for detection and response, and ultimately protect public health. Results from our study further support the value of this approach.

## Acknowledgements

This work was supported by the Lawrence Livermore National Laboratory’s Laboratory Derived Research & Development program. The residual samples used in this study were obtained from the original community surveillance project CalSRVSS which was funded in part and by the Centers for Disease Control and Prevention Epidemiology and Laboratory Capacity for Community Surveillance (grant 6 Nu50CK000539).

## Author Contributions

RSK, ACM, CRK and CJ contributed to the writing of the of the draft manuscript. RSK, CRK, ACM and JBT analyzed and interpreted the data. CJ, DAW, CM and SM acquired the samples. JBT, ACM, MB, CJ, DAW, CM and SM designed the study and experiments. ACM, JBT, DAW, SM and CM performed experiments. CJ and DAW contributed to project management. All authors contributed to reviewing and editing the final manuscript.

## Data availability

The de-identified untargeted metagenomic and TWIST-enriched Illumina sequencing datasets are publicly available in the NCBI Sequencing Read Archive under BioProject PRJNA1296491.

## Notes

### Competing Interest Statement

The authors have declared no competing interest.

### Funding Statement

This work was supported by the Lawrence Livermore National Laboratory Derived Research & Development program. The residual samples used in this study were obtained from the original community surveillance project CalSRVSS which was funded in part and by the Centers for Disease Control and Prevention Epidemiology and Laboratory Capacity for Community Surveillance (grant 6 Nu50CK000539).

### Author Declarations

Ethics committee/IRB of California Department of Public Health Human Services Agency Committee waived ethical approval for this work.

