## Supplementary Figures for "Metagenomic sequencing identifies potential respiratory pathogens in PCR-negative subset of surveillance samples"

### Supplementary Information

Tables:

Supplementary Table S1

Figures:

Supplementary Figure S1

Supplementary Figure S2

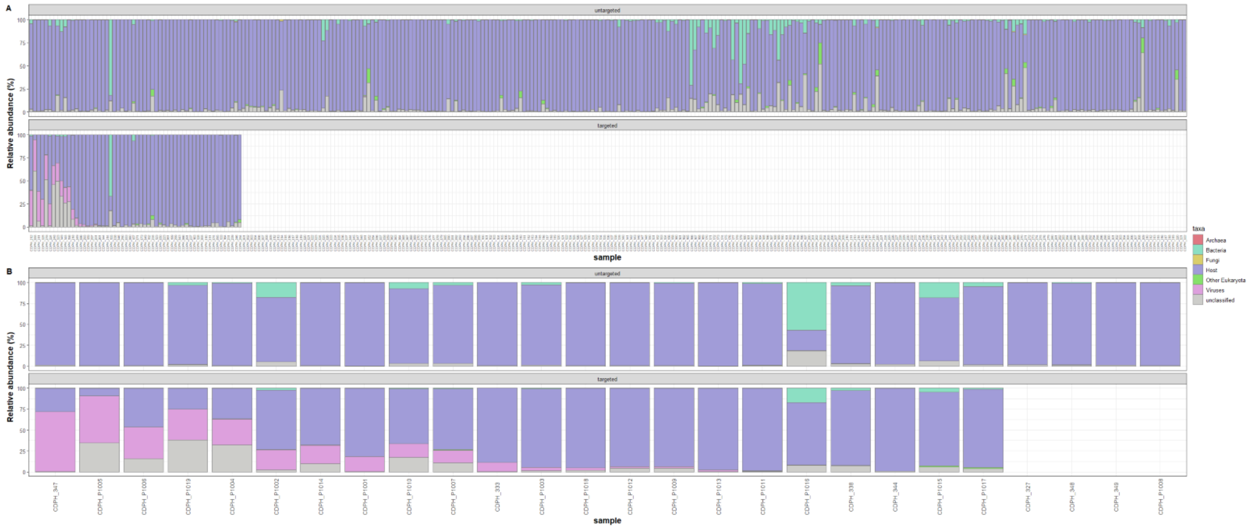


**Supplementary Figure S1. The percentage of reads assigned to host or to a given microbial group.** Samples are divided into (A) PCR-negative and (B) PCR-positive sets, and results of untargeted sequencing (top panel) and targeted sequencing (bottom panel) are shown for each. Samples arranged according to viral abundance in targeted/enriched samples as classified by Recentrifuge.

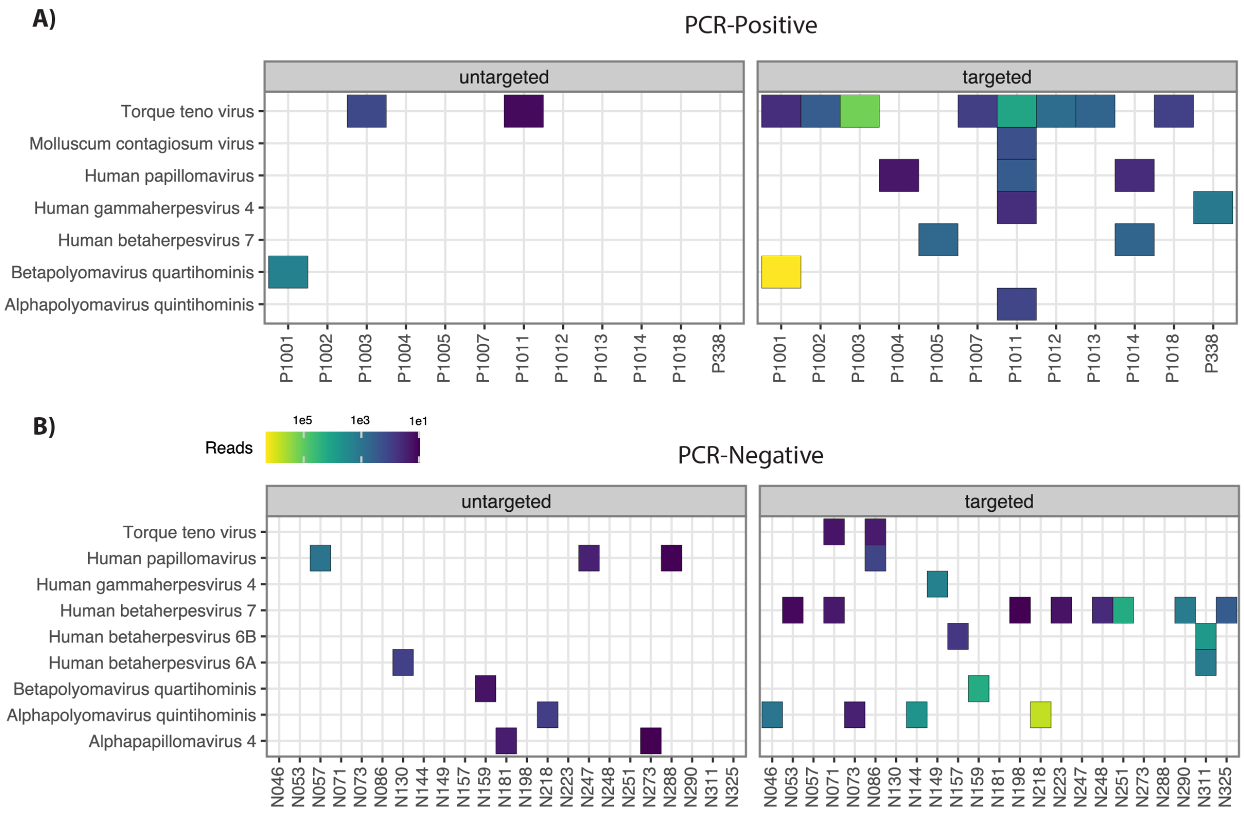


**Supplementary Figure S2. Detection of non-respiratory viruses.** Read count results for untargeted and targeted sequencing are displayed for (A) PCR-negative samples and (B) PCR positive samples. These viruses include polyomaviruses, papillomaviruses, various herpes viruses, and Torque teno virus, which are commonly carried in healthy individuals. We reported the non-respiratory viruses detected by Centrifuge with ≥10 reads per sample.
